# Comparative Survival Analysis of Immunomodulatory Therapy for COVID-19 ‘Cytokine Storm’: A Retrospective Observational Cohort Study

**DOI:** 10.1101/2020.06.16.20126714

**Authors:** Sonali Narain, Dimitre G. Stefanov, Alice S. Chau, Andrew G. Weber, Galina Marder, Blanka Kaplan, Prashant Malhotra, Ona Bloom, Audrey Liu, Martin L. Lesser, Negin Hajizadeh, Northwell COVID-19 Research Consortium

## Abstract

**Background:** Cytokine storm is a marker of COVID-19 illness severity and increased mortality. Immunomodulatory treatments have been repurposed to improve mortality outcomes.

**Methods:** We conducted a retrospective analysis of electronic health records across the Northwell Health system. COVID-19 patients hospitalized between March 1, 2020 and April 15, 2020, were included. Cytokine storm was defined by inflammatory markers: ferritin >700ng/mL, C-reactive protein >30mg/dL, or lactate dehydrogenase >300U/L. Patients were subdivided into six groups -no immunomodulatory treatment (standard of care) and five groups that received either corticosteroids, anti-interleukin 6 (IL-6) antibody (tocilizumab) or anti-IL-1 therapy (anakinra) alone or in combination with corticosteroids. The primary outcome was hospital mortality.

**Results:** There were 3,098 patients who met inclusion criteria. The most common comorbidities were hypertension (40-56%), diabetes (32-43%) and cardiovascular disease (2-15%). Patients most frequently met criteria with high lactate dehydrogenase (74.8%) alone, or in combination, followed by ferritin (71.4%) and C-reactive protein (9.4%). More than 80% of patients had an elevated D-dimer. Patients treated with a combination of tocilizumab and corticosteroids (Hazard Ratio [HR]: 0.459, 95% Confidence Interval [CI]: 0.295-0.714; p<0.0001) or corticosteroids alone (HR: 0.696, 95% CI: 0.512-0.946; p=0.01) had improved hospital survival compared to standard of care. Corticosteroids and tocilizumab was associated with increased survival when compared to corticosteroids and anakinra (HR: 0.612, 95% CI: 0.391-0.958; p-value=0.02).

**Conclusions:** When compared to standard of care, corticosteroid and tocilizumab used in combination, or corticosteroids alone, was associated with reduced hospital mortality for patients with COVID-19 cytokine storm.

## Introduction

In March 2020, New York City and its metropolitan area became the epicenter for coronavirus disease 2019 (COVID-19) in the United States, with over 250,000 cases and greater than 17,000 deaths by early May 2020.^1^ Throughout this outbreak, physicians and scientists have struggled to understand the pathogenesis and clinical course of this infection. Early retrospective data from China and Italy showed increased mortality in those with elevated inflammatory markers, such as ferritin, C-reactive protein (CRP), lactate dehydrogenase (LDH), interleukin 6 (IL-6) and D-dimer.^2^ Uncontrolled and unabated cytokine release and a hyperinflammatory response termed as COVID-19 “cytokine storm” (CCS), was described as a major determinant of poor survival.^3^

Limited data existed to guide clinical decision-making in the absence of FDA-approved COVID-19 specific therapies. Faced with rapidly increasing rates of infection and hospitalizations, physicians repurposed immunomodulatory treatments in an attempt to curtail morbidity and mortality. Although initial reports discouraged the use of corticosteroids, later publications suggested survival benefits.^2,4,5^ Small retrospective studies reported improved outcomes in CCS by using anti-IL-6 (i.e., tocilizumab) and anti-IL-1 therapies (i.e., anakinra)^6-8^ that are commonly used for inflammatory conditions such as cytokine release syndrome and macrophage-activation syndrome. Further evidence supporting the use of anti-IL-1 was based on previous reports of improved survival in a subgroup of patients with sepsis and hyperferritinemia.^9^

Within Northwell Health, the largest private nonprofit health system in New York state, a multidisciplinary committee consisting of pulmonology, infectious disease, immunology and rheumatology specialists was formed to create COVID-19 treatment protocols. This included the identification of CCS and options for treatment with corticosteroids, tocilizumab and anakinra as potential immunomodulatory therapies based on the available literature.^2,10,11^ Due to the rapidly evolving data and surge of patients in a short timeframe, there was wide variation in the use of these drugs across the health system. In this retrospective study, we leverage this natural experiment to compare outcomes for CCS patients who received different combinations of these immunomodulatory drugs.

## Methods

### Study Population

We retrospectively analyzed electronic health record data of patients admitted to the 12 hospitals and emergency departments within the Northwell Health system between March 1, 2020 and April 15, 2020. The Institutional Review Board for the Feinstein Institutes of Medical Research at Northwell Health approved this study as minimal-risk research and waived the requirement for informed consent. COVID-19 positivity was determined by polymerase chain reaction testing of nasopharyngeal swabs. Among patients who tested positive, we identified those who met our CCS criteria: ferritin > 700ng/mL^11^ or CRP > 30mg/dL^2,10^ or LDH > 300U/L^2^ (Figure 1). The time at which a patient was first identified as meeting this definition was labeled as “T_0_”. Patients under age 18 were excluded.

**Figure 1.**
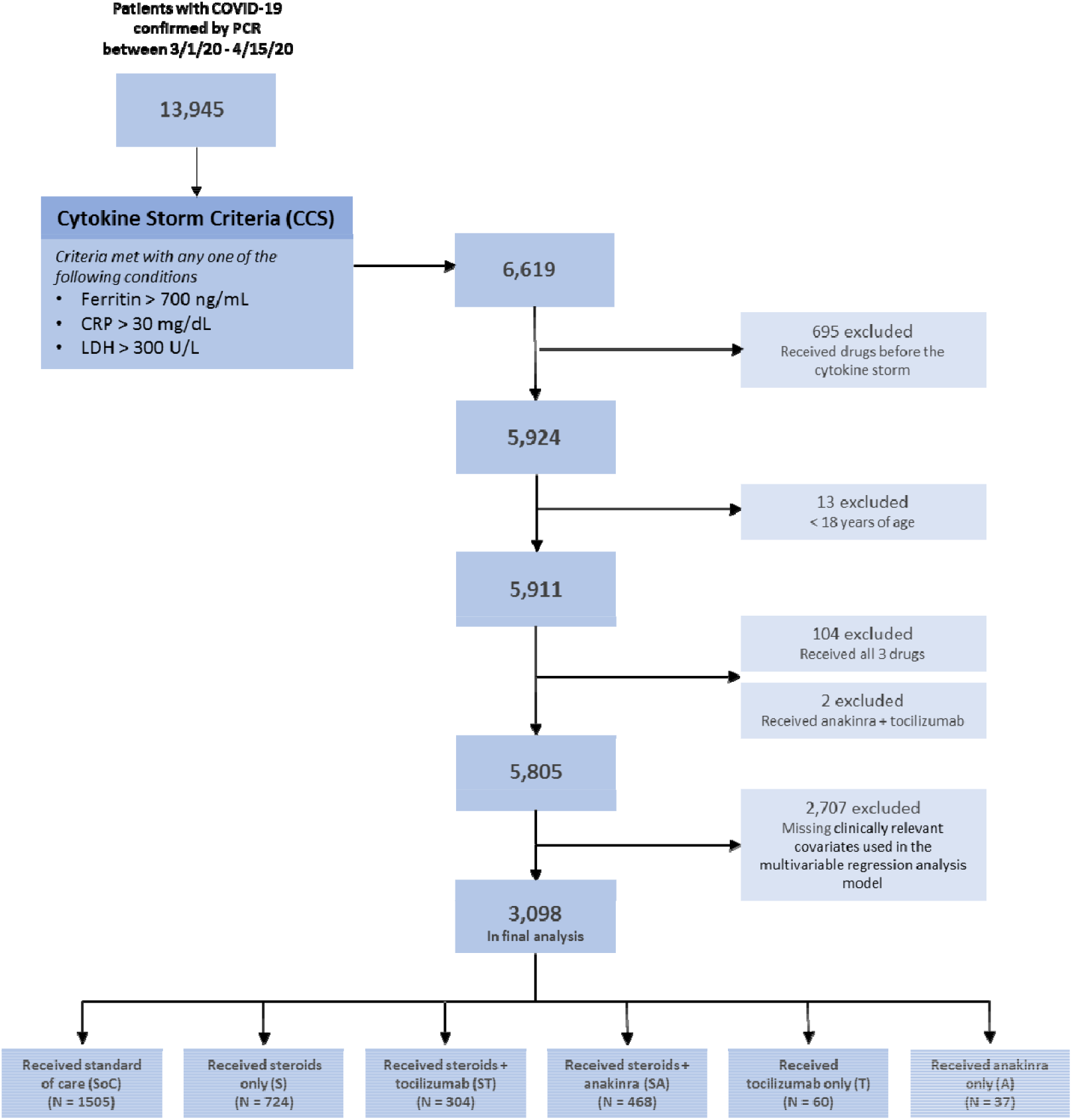
Consort Diagram of Retrospective Cohort Study. Consort diagram showing selection of patients, inclusion criteria, and exclusion criteria applied to form the final cohort of 3,098 patients. Exclusion criteria included receiving any of the immunomodulatory drugs prior to the diagnosis of cytokine storm, age <18 years, having received all 3 study drugs, having received the combination of anakinra and tocilizumab, or missing clinically relevant covariates. 3,098 patients remained in the final analysis. PCR = polymerase chain reaction testing

### Group definition

There were six groups identified based on whether they received any of the pre-defined immunomodulatory drugs. One group consisted of those who received none of the medications, labeled as standard of care (SoC). Five treatment groups received varying combinations of the three immunomodulatory drugs: corticosteroids only (S), corticosteroids and tocilizumab (ST), corticosteroids and anakinra (SA), tocilizumab only (T) and, anakinra only (A). In the timeframe of this analysis, almost all COVID-19 patients received hydroxychloroquine as part of institutional protocols.

### Statistical methods

#### Primary objective

To compare in-hospital mortality among COVID-19 patients with CCS who received combinations of immunomodulatory treatments versus SoC.

#### Covariates

Potentially confounding variables (covariates) were included in the multivariable model based on clinical experience and the COVID-19 literature. These included demographic data such as age, gender, race/ethnicity and, insurance status. Comorbidities examined included chronic lung disease (i.e., smoking history, asthma, chronic obstructive pulmonary disease), cardiovascular disease, hypertension, diabetes, renal disease, hemodialysis, liver disease, cancer, autoimmune disease and, the Charlson comorbidity index (CCI). Laboratory data included CRP, ferritin, D-dimer, hemoglobin, platelet count, serum sodium, serum transaminases and, neutrophil to lymphocyte ratio (NLR). We also included disease severity surrogates, such as use of invasive mechanical ventilation (IMV; within 24 hours of T_0_) and, vasopressor use (within 24 hours of T_0_). Body mass index could not be included in the analysis due to a large amount of missing data.

### Statistical analyses

Treatment groups were compared using demographic variables, comorbidities and baseline lab values using the χ^2^, Fisher’s exact or Kruskal-Wallis tests, as appropriate. Categorical variables were summarized using percentages, means and, standard deviations. Continuous variables were summarized using medians with 25-75^th^ percentiles (PCT). Labs considered clinically important were included in the Cox regression analysis except for LDH, which was excluded due to a large number of missing values. Baseline laboratory values in this study were defined as the value closest to T_0_ within the 96 hours prior to T_0_. Exceptions were for CRP, ferritin and D-dimer, which were defined as within 96 hours prior to T_0_ and up to 12 hours after T_0_, due to laboratory ordering practices. Patient survival was calculated from T_0_ to the time of in-hospital death. Data from patients discharged from the hospital or remaining in the hospital on April 24, 2020 were considered censored.

In-hospital patient survival was compared between treatment groups for patients who had all demographics and comorbidity data and laboratory variables available in the pre-specified windows (complete covariate data). To determine the risk of selection bias, we compared survival between patients--with and without complete covariate data--using the Kaplan-Meier product limit method and, the log-rank test. The two groups were not significantly different (Supplemental Figure 1). Patient survival was compared between treatment groups using the Cox regression model, adjusting for all covariates outlined above. The proportional hazards assumption was assessed and, deemed acceptable. Tukey’s adjustment for multiple comparisons was used to account for the 15 pairwise tests resulting from the six groups.

The final model included all clinically important covariates regardless of their statistical significance (the “full model”). To test the robustness of the model, we considered two other analytical approaches. First, we used backward selection to find a parsimonious set of covariates which were used to adjust the model. Next, we excluded the two treatment arms with relatively small sample sizes (those who received either A or T). In both tests, the results remained consistent with those of the full model.

SAS 9.4 (SAS Institute, Cary, NC) was used for the statistical analysis. Results were considered statistically significant if *p*<0.05.

## Results

### Patient characteristics

Of the 13,945 patients with COVID-19, seen in emergency departments or admitted to hospitals within the Northwell Health system during the study period, 6,619 (45.7%) patients met at least one criterion for the definition of CCS. Of these, 3,098 patients were included in the final analysis (Figure 1).

Demographic characteristics and distribution of covariates across groups are reported in Table 1. There were twice as many males as females. A significant difference in the racial distribution across treatment groups was noted, with more black and multiracial patients in the A group. Most of the cohort had never smoked. The most common comorbidities across groups were: hypertension (40-56%), diabetes (32-43%), cardiovascular disease (2-15%), chronic kidney disease (5-11%), cancer (5-13%) and asthma (2-8%). Only 2% of patients were on hemodialysis prior to T_0_. Approximately, 40% of the patients in the cohort had low predicted 10-year survival rate based on CCI (≥5). There were more patients with low CCI (0) in the SoC group as compared to other treatment groups. Approximately, 6% of patients were on IMV and 4% on vasopressors at T_0_. More than 80% of the patients who met criteria for CCS had an elevated D-dimer, of which ∼20% had levels greater than five times the upper limit of normal.

**Table 1:**
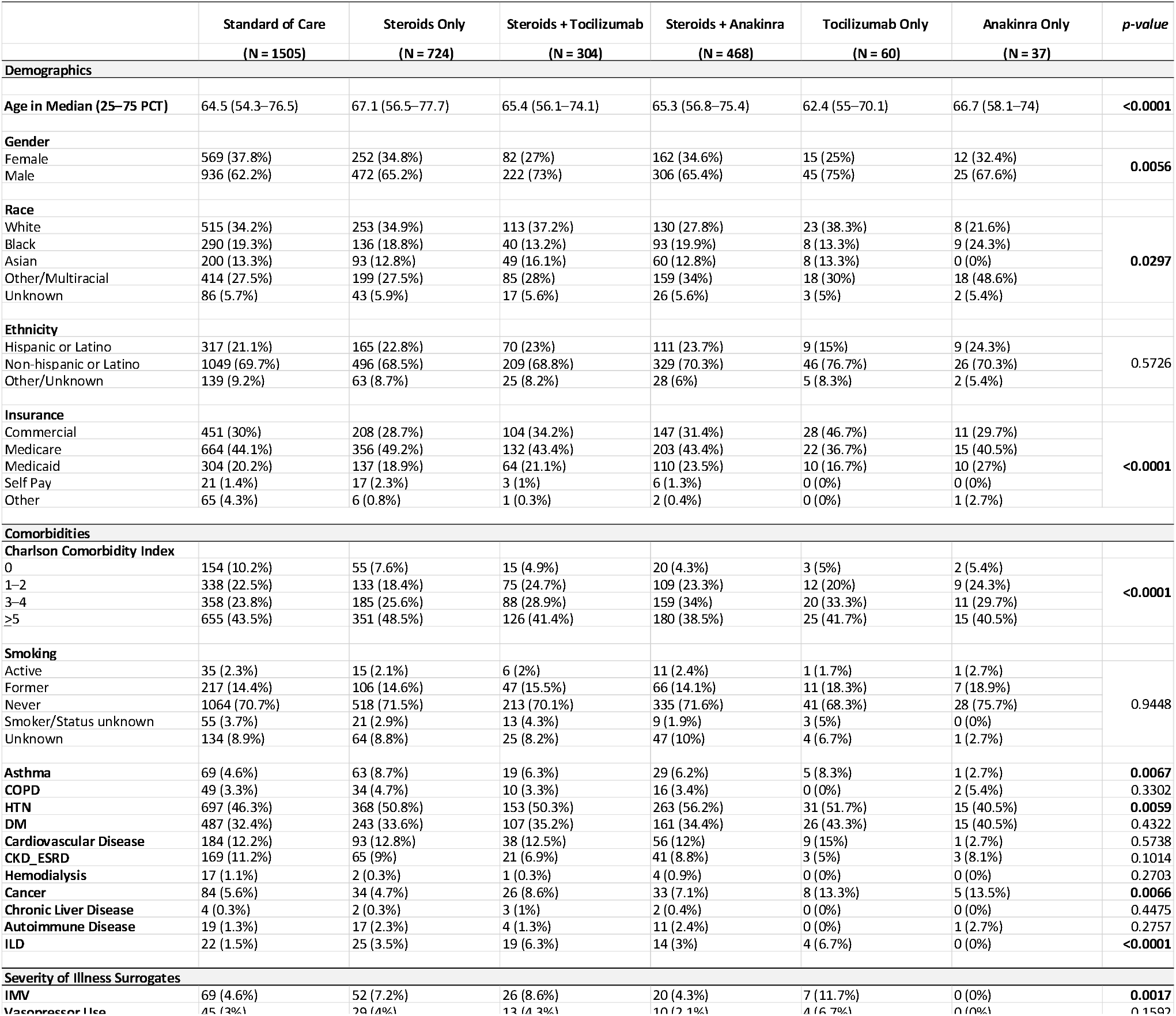
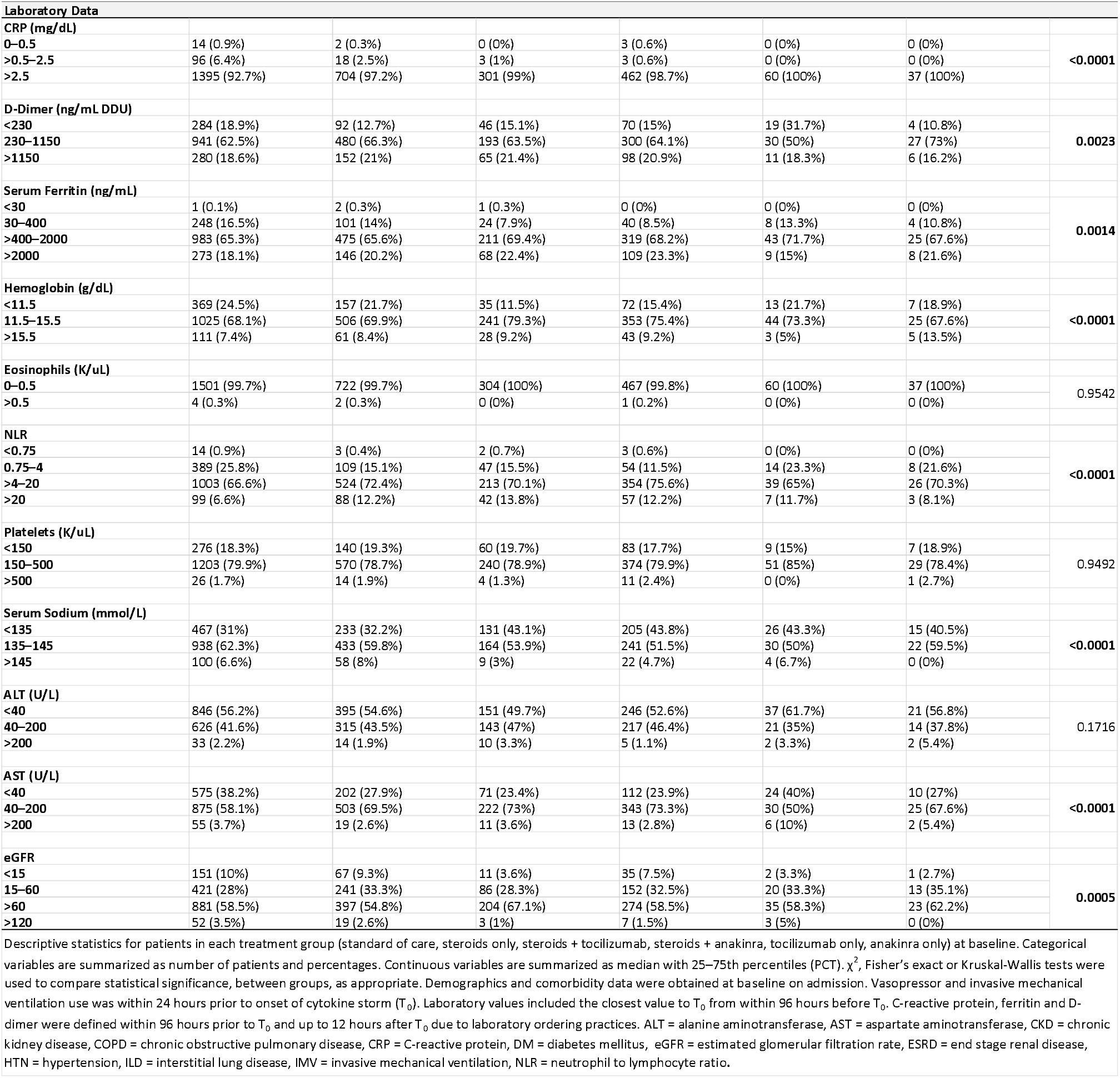
Patient Demographics.

The most common criterion met for CCS definition was high LDH, which was found in 74.8% of patients, either alone or in combination with other criteria, followed by ferritin (71.4%) and CRP (9.4%). The definition of CCS was met by only one criterion in 49.5% of patients, by two criteria in 45.4% and by three criteria in 5.1% of patients. The distribution of CRP, ferritin and LDH is provided in Supplemental Figure 2. There was a statistically significant variation between treatment arms with respect to CRP, ferritin and LDH (*p*<0.0001).

**Figure 2:**
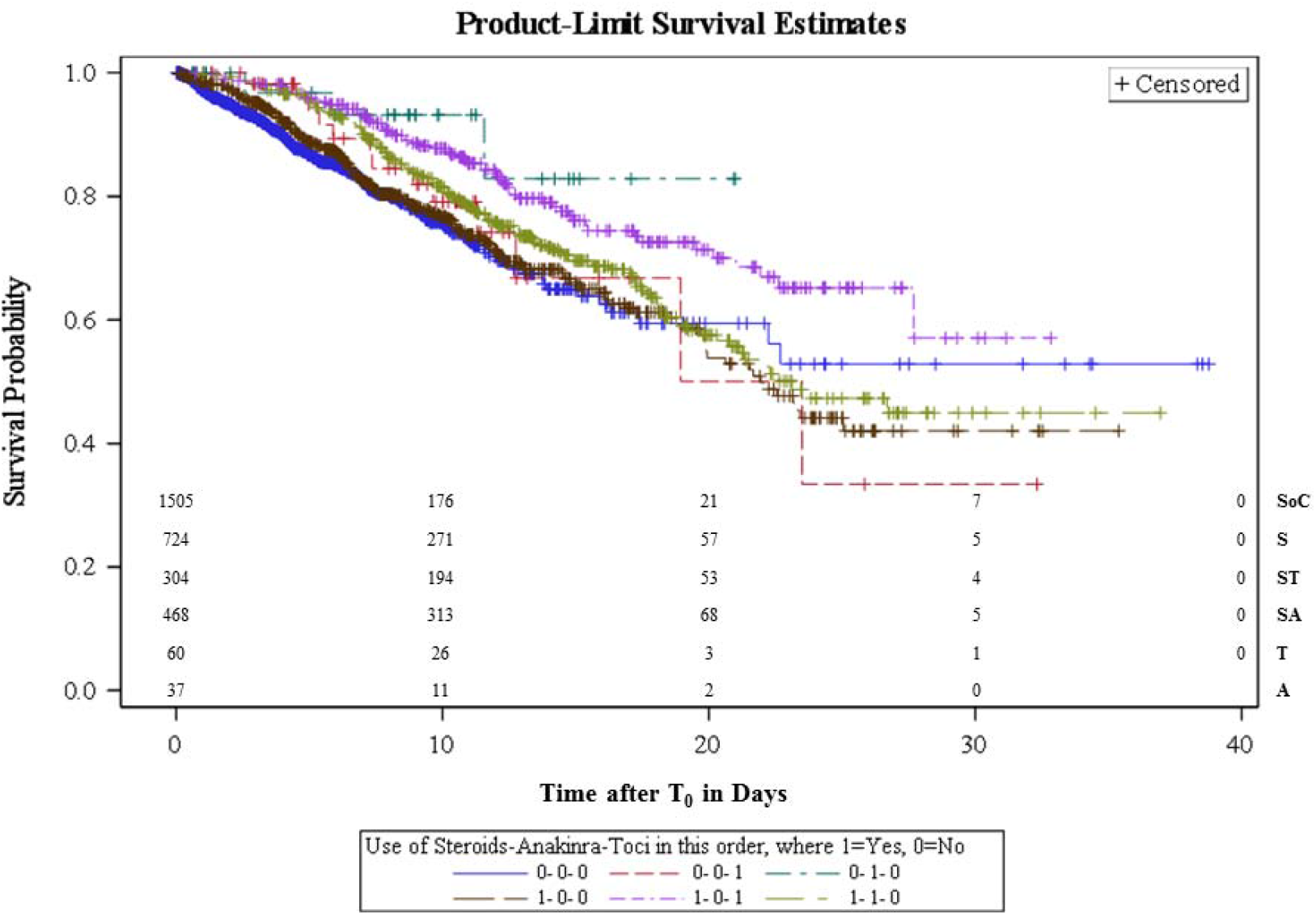
Kaplan-Meier plots for treatment groups (unadjusted for covariates) This figure represents the unadjusted Kaplan Meier plots for treatment groups with number of subjects at risk. The treatment groups are as follows: SoC (0-0-0) = standard of care, S (1-0-0) = steroid only, T (0-0-1) = tocilizumab only, A (0-1-0) = anakinra only, ST (1-0-1) = steroids + tocilizumab, SA (1-1-0) = steroids + anakinra.

Kaplan-Meier plots for treatment groups (unadjusted for covariates) and model based Kaplan-Meier survival estimates for treatment groups (adjusted for covariates) are presented in Figures 2 and 3 respectively. A Cox proportional hazards model was used to compare treatment groups adjusting for clinically important variables. In this model, covariates that were statistically significantly associated with increased mortality were older age, unknown smoking status and self-pay insurance (Table 2). Higher mortality was associated with presence of interstitial lung disease (ILD), cardiovascular disease and, the need for IMV at T_0_. Elevated D-dimer and NLR were also associated with higher mortality. Hypertension was associated with lower mortality.

**Table 2:** Cox Regression Models.

| TABLE 2: Hazard Ratios with 95% Confidence Intervals for Cox Regression Model |  |  |  |  |  |  |  |  |
| --- | --- | --- | --- | --- | --- | --- | --- | --- |
|  | Hazard Ratio (95% Confidence Limits) | p-value |  | Hazard Ratio (95% Confidence Limits) | p-value |  | Hazard Ratio (95% Confidence Limits) | p-value |
| **Treatment Groups\*** |  |  | **Laboratory parameters** |  |  | **Comorbidities** |  |  |
| *Standard of Care* | reference |  | **Eosinophils (K/uL)** |  |  | **Smoking status** |  |  |
| *Steroids only* | 0.696 (0.563 - 0.860) | 0.0008 | 0 - 0.5 | reference |  | Never | reference |  |
| *Steroids + Tocilizumab* | 0.459 (0.399 - 0.622) | <0.0001 | > 0.5 | 0.725 (0.099 - 5.287) | 0.7511 | Active | 1.233 (0.668 - 2.277) | 0.5035 |
| *Steroids + Anakinra* | 0.750 (0.596 - 0.944) | 0.0143 | **Platelets (K/uL)** |  |  | Former | 1.002 (0.791 - 1.271) | 0.9838 |
| *Tocilizumab only* | 0.718 (0.403 - 1.280) | 0.2615 | 150 - 500 | reference |  | Smoker/Current Status | 1.427 (0.982 - 2.074) | 0.0621 |
| *Anakinra only* | 0.404 (0.128 - 1.278) | 0.1230 | < 150 | 1.222 (1.008 - 1.482) | **0.0415** | Unknown | 2.702 (2.149 - 3.398) | **< 0.0001** |
|  |  |  | > 500 | 1.030 (0.567 - 1.871) | 0.9231 |  |  |  |
| **Demographics** |  |  | **Hemoglobin (g/dL)** |  |  | **Charlson comorbidity Index** |  |  |
| **Age** | 1.034 (1.023 - 1.044) | **< 0.0001** | 11.5 - 15.5 | reference |  | 0 | reference |  |
|  |  |  | < 11.5 | 1.162 (0.952 - 1.418) | 0.1395 | 1 - 2 | 1.066 (0.489 - 2.325) | 0.8725 |
|  |  |  | > 15.5 | 1.024 (0.755 - 1.388) | 0.8798 | 3 - 4 | 1.143 (0.523 - 2.499) | 0.7369 |
| **Gender** |  |  | **eGFR** |  |  | > 5 | 1.368 (0.600 - 3.119) | 0.4569 |
| Female | reference |  | 60 - 120 | reference |  |  |  |  |
| Male | 1.062 (0.883 - 1.276) | 0.5240 | < 15 | 1.992 (1.448 - 2.741) | **< 0.0001** | **Asthma** |  |  |
|  |  |  | 15 - 60 | 1.704 (1.391 - 2.087) | **< 0.0001** | No | reference |  |
|  |  |  | > 120 | 1.328 (0.557 - 3.166) | 0.5216 | Yes | 1.331 (0.885 - 2.001) | 0.1691 |
| **Race** |  |  | **AST (u/L)** |  |  | **COPD** |  |  |
| White | reference |  | 0 - 40 | reference |  | No | reference |  |
| Asian | 0.973 (0.755 - 1.254) | 0.8329 | > 40 | 1.281 (1.034 - 1.587) | **0.0234** | Yes | 1.341 (0.929 - 1.938) | 0.1175 |
| Black | 0.797 (0.625 - 1.018) | 0.0691 | > 200 | 1.036 (0.632 - 1.700) | 0.8880 | **Chronic Liver Disorder** |  |  |
| Other/Multiracial | 0.865 (0.663 - 1.129) | 0.2857 | **ALT (u/L)** |  |  | No | reference |  |
| Unknown | 0.978 (0.571 - 1.676) | 0.9367 | 0 - 40 | reference |  | Yes | 0.740 (0.180 - 3.035) | 0.6760 |
| **Ethnicity** |  |  | > 40 | 0.907 (0.742 - 1.109) | 0.3421 | **DM** |  |  |
| Not Hispanic or Latino | reference |  | > 200 | 1.473 (0.823 - 2.634) | 0.1920 | No | reference |  |
| Hispanic or Latino | 0.880 (0.668 - 1.160) | 0.3656 | **Sodium (mmol/L)** |  |  | Yes | 1.024 (0.846 - 1.240) | 0.8090 |
| Other/Unknown | 0.781 (0.472 - 1.292) | 0.3354 | 135 - 145 | reference |  | **HTN** |  |  |
|  |  |  | < 135 | 1.153 (0.956 - 1.389) | 0.1367 | No | reference |  |
| **Insurance** |  |  | > 145 | 1.284 (0.985 - 1.674) | 0.0651 | Yes | 0.749 (0.629 - 0.892) | **0.0012** |
| Commercial | reference |  | **Ferritin (ng/mL)** |  |  | **ILD** |  |  |
| Medicaid | 1.109 (0.810 - 1.519) | 0.5183 | 30 - 400 | reference |  | No | reference |  |
| Medicare | 1.058 (0.818 - 1.369) | 0.6657 | < 30 | 3.081 (0.404 - 23.479) | 0.2775 | Yes | 2.272 (1.630 - 3.169) | **< 0.0001** |
| Other | 1.077 (0.319 - 3.640) | 0.9049 | > 400 | 1.000 (0.764 - 1.307) | 0.9975 | **Autoimmune Disorder** |  |  |
| Self Pay | 3.275 (1.825 - 5.878) | **< 0.0001** | > 2000 | 1.120 (0.818 - 1.532) | 0.4793 | No | reference |  |
|  |  |  | **CRP (mg/dL)** |  |  | Yes | 1.146 (0.605 - 2.168) | 0.6765 |
| **Disease severity indices** |  |  | 0 - 0.5 | reference |  | **Cardiovascular disease** |  |  |
| **Mechanical ventilation** |  |  | > 0.5 | 1.684 (0.210 - 13.493) | 0.6233 | No | reference |  |
| No | reference |  | > 2.5 | 2.316 (0.311 - 17.259) | 0.4124 | Yes | 1.286 (1.026 - 1.612) | **0.0293** |
| Yes | 1.472 (1.075 - 2.017) | **0.0160** | **D-Dimer (ng/mL DDU)** |  |  | **CKD** |  |  |
| **On vasopressors** |  |  | 0 - 230 | reference |  | No | reference |  |
| No | reference |  | > 230 | 1.693 (1.216 - 2.357) | 0.0018 | Yes | 0.779 (0.579 - 1.047) | 0.0976 |
| Yes | 0.891 (0.616 - 1.287) | 0.5375 | > 1150 | 2.097 (1.468 - 2.995) | **< 0.0001** | **Cancer** |  |  |
|  |  |  | **NLR** |  |  | No | reference |  |
|  |  |  | 0.75 - 4 | reference |  | Yes | 1.332 (0.979 - 1.812) | 0.0683 |
|  |  |  | < 0.75 | 1.709 (0.769 - 3.803) | 0.1887 | **Hemodialysis** |  |  |
|  |  |  | > 4 | 1.368 (1.060 - 1.766) | 0.0161 | No | reference |  |
|  |  |  | > 20 | 1.117 (0.805 - 1.549) | 0.5079 | Yes | 0.979 (0.387 - 2.481) | 0.9648 |

**Figure 3:**
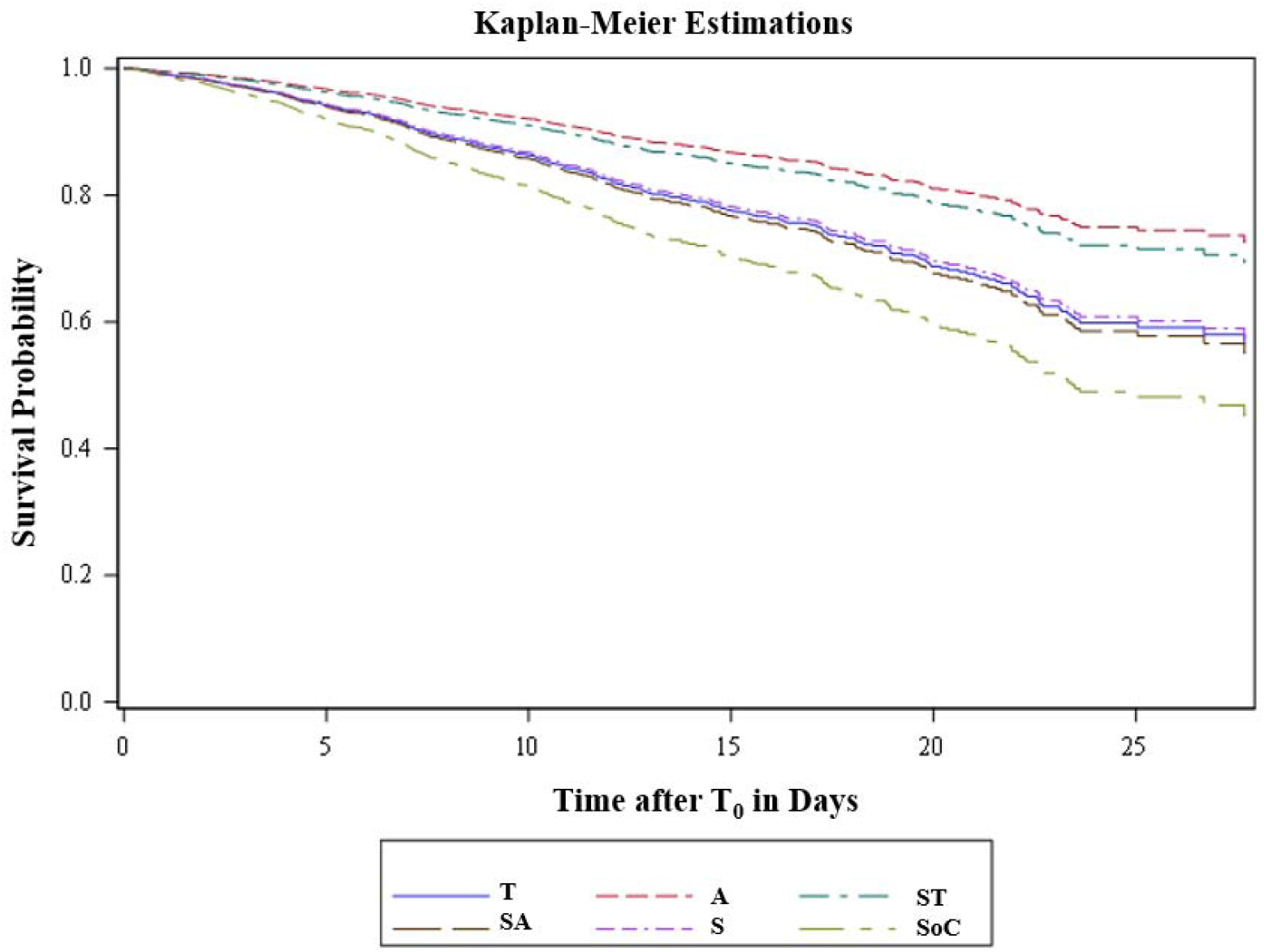
Kaplan-Meier Model-Based Survival Estimates. The graph represents the model-based survival estimates for each treatment groups. Covariates included in the model are categories with highest frequencies and mean age of 65.2 years. Specifically, laboratory values were set to normal level categories with the following exceptions: above normal categories for lactate dehydrogenase, serum ferritin, D-dimer, neutrophil to lymphocyte ratio; very high category for C-reactive protein. SoC = standard of care, S = steroid only, T = tocilizumab only, A = anakinra only, ST = steroids + tocilizumab, SA = steroids + anakinra.

**Figure 4:**
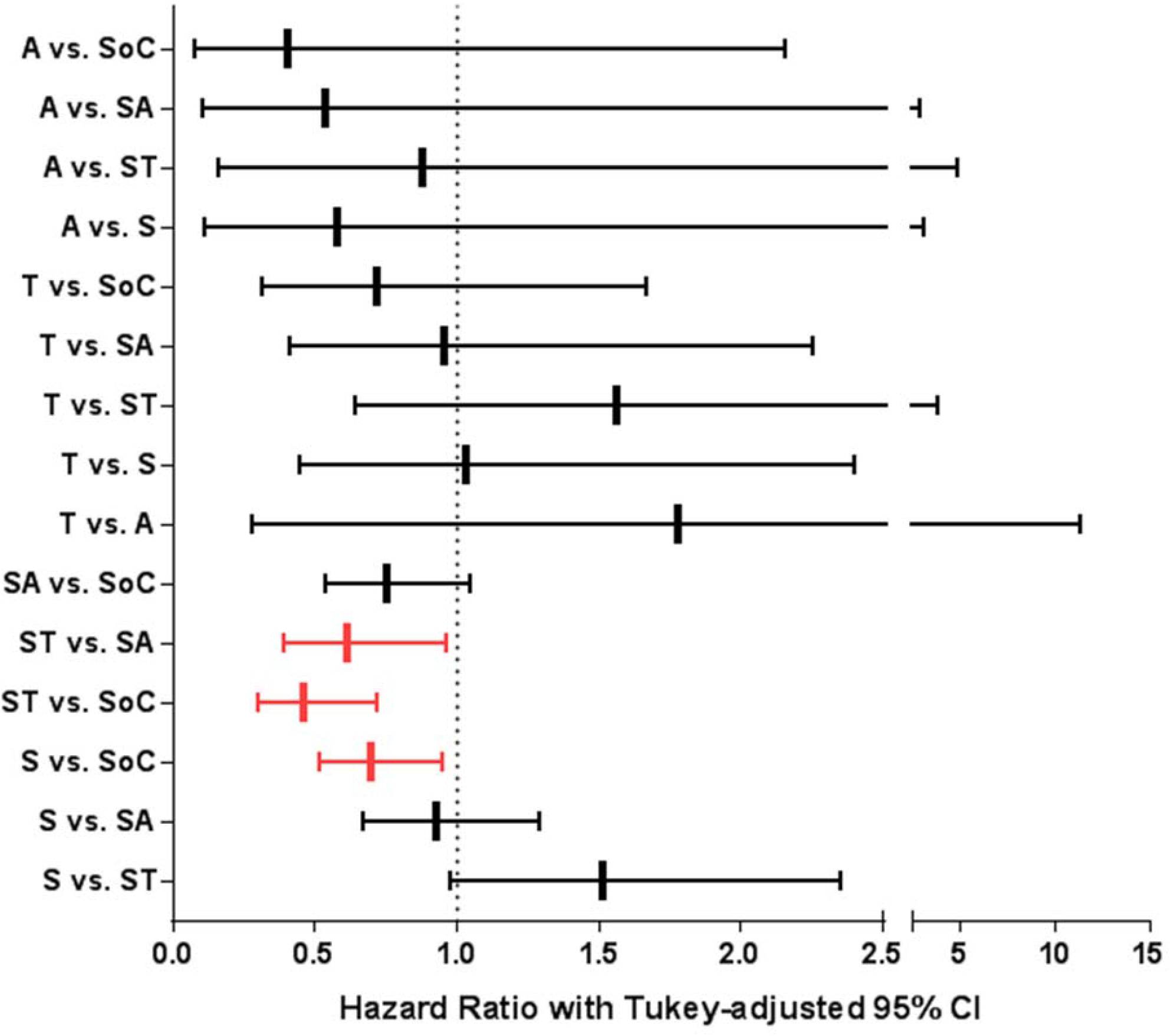
Hazard Ratios for Treatment Differences Using Tukey’s Adjustment for Multiple Comparisons. The figure represents pairwise comparisons for all treatment groups with Tukey’s adjustment for multiple comparison. The groups are as follows: SoC = standard of care, S = steroid only, ST = steroids + tocilizumab, SA = steroids + anakinra, T = tocilizumab only, A = anakinra only. Groups in red are statistically significant.

Pairwise comparisons between treatment groups are presented in Figure 3. Patients in the ST and S groups had significantly improved survival compared to the SoC group (ST vs. SoC Hazard Ratio (HR): 0.459, 95% confidence interval (CI): 0.295-0.714; *p*<0.0001; S vs. SoC HR: 0.696, 95% CI: 0.512-0.946; *p*=0.01). However, patients in the SA or A groups did not have significantly different survival when compared to SoC. When comparing the treatment groups with each other, the ST group appeared to have improved outcomes compared to SA (HR: 0.612, 95% CI: 0.391-0.958; p=0.02). No other significant differences were seen between the other treatment groups.

At T_0_, in patients receiving only one of the three treatments, corticosteroids were started earlier (median: 19.72hrs; 25^th^-75^th^ PCT 6.06-57.94) than either tocilizumab (median: 52.66hrs; 25^th^-75^th^ PCT 25.30-98.78) vs. or, anakinra (median: 46.95hrs; 25^th^-75^th^ PCT 20.87-96.35). In both groups that received combination therapy with corticosteroids, corticosteroids were started before the second drug and, at a similar interval from T_0_ (Supplemental Figure 3). The time from T_0_ to tocilizumab dosing was comparable when used alone (median: 52.66hrs; 25^th^-75^th^ PCT 25.30-98.78) or, in combination with corticosteroids (median: 49.50hrs; 25^th^-75^th^ PCT 18.62-100.28). Anakinra alone was given earlier (median: 46.95hrs; 25^th^-75^th^ PCT 20.87-96.35) than anakinra in the SA group (median: 70.17hrs; 25^th^-75^th^ PCT 30.50-119.07).

## Discussion

This large retrospective observational study leverages natural heterogeneity in practice patterns for COVID-19 CCS patients. We describe hospital survival outcomes in patients receiving different combinations of immunomodulatory therapy with careful consideration of potential confounders available in the electronic health records. Our findings suggest that tocilizumab and corticosteroids used together or corticosteroids used alone were associated with lower mortality as compared to SoC. This association remained after controlling for covariates that influence mortality in COVID-19.

Age was associated with increased mortality regardless of treatment group--consistent with other COVID-19 survival analyses. Self-pay insurance status (<2% of patients) was associated with increased mortality, which may be explained by socioeconomic disadvantages in this group. We speculate that self-pay patients may have presented to the hospital later in disease course. For surrogates of illness severity, the need for IMV prior to T_0_ was associated with increased mortality, while the need for vasopressors was not.

Prior diagnoses of ILD, cardiovascular disease and renal dysfunction were associated with increased mortality, consistent with existing literature.^12^ Surprisingly, those with comorbid hypertension had lower mortality which is contradictory to other reports.^13,14^ Interestingly, one study suggested that use of angiotensin-converting enzyme inhibitors or angiotensin II receptor blockers via renin-angiotensin pathway modulation may confer a protective effect in the setting of CCS.^15^ Our analysis did not include consideration of home medications. Alternatively, adjustments for covariates in our model may have uncovered an association between hypertension and COVID-19 outcomes which can be further investigated.

High D-dimer was significantly associated with in-hospital mortality. This is consistent with mounting evidence that elevated D-dimer is associated with worse outcomes^16^ and predicts a higher chance of requiring intensive care unit (ICU) admission and increased 28-day mortality.^4,17,18^ Zhou *et al*., showed a steep rise in D-dimer by day 10 of illness that separated non-survivors.^17^ We also found thrombocytopenia to be associated with higher mortality. Both thrombocytopenia and elevated D-dimer reflect the known coagulopathy in COVID-19.

IL-6 is an important mediator of inflammation that plays an essential role in host response to viral infection. High IL-6 levels were observed in patients with severe COVID-19.^2^ Therefore, tocilizumab was proposed early in the COVID-19 pandemic as a potential treatment for those with CCS. In two small case series studying severe COVID-19, improvements in clinical and mortality outcomes were reported with the use of intravenous^19^ and subcutaneous tocilizumab.^20^ Toniati *et al*. prospectively treated 100 patients with intravenous tocilizumab and reported 20% mortality, despite 43% of their patients being in the ICU at the time of intervention.^6^ Conversely, Colaneri *et al*. reported a retrospective analysis of 21 patients treated with IV tocilizumab and saw no reduction of ICU admissions or mortality.^21^ In our cohort, patients who received ST were more likely to survive compared to SoC as well as the SA arm. Tocilizumab alone did not improve survival.

Although corticosteroids are used in the treatment of hyperinflammatory syndromes and acute respiratory distress syndrome,^22^ their use in viral infections and severe acute respiratory syndrome coronavirus 2 (SARS-CoV-2) is controversial and not currently recommended by the World Health Organization.^23^ Early in the COVID-19 pandemic, Wang *et al*. reported an absence of a protective effect from corticosteroids.^4^ However, Wu *et al*. reported a survival benefit of ∼15% in critically ill patients. Lower mortality was reported in the methylprednisolone group (46%) versus no treatment (61.8%).^2^ While there was concern that use of corticosteroids may prolong viral shedding, Fang *et al*. reported that viral clearance was not affected when used later in the disease course.^5^ Our results suggest a benefit from corticosteroids when used alone or in combination with tocilizumab.

Anti-IL-1 therapy has been an attractive choice in the treatment of COVID-19 due to its short half-life, safety and tolerability profile. IL-1β has been implicated in lung inflammation, fibrosis^24^ and, indirectly, with activation of the inflammatory cascade.^25-28^ A study examining cytokine kinetics during COVID-19 showed an IL-1 peak prior to the apex of respiratory distress and the surge of other inflammatory cytokines.^29^ A study of anakinra in sepsis demonstrated improved survival in a subset of patients with hyperferritinemia and hepatobiliary dysfunction^9^ as compared to placebo.^30^

Small studies report improvement in clinical outcomes with use of anakinra in COVID-19.^7,8^ Cavalli *et al*. evaluated 36 hospitalized non-ICU patients with CCS and observed improvement in respiratory function, inflammatory markers and intubation avoidance in 72% of patients receiving high dose intravenous anakinra as compared to low dose or SoC.^7^ In this study of patients treated with anakinra, either alone or in combination with corticosteroids, no mortality benefit was seen with either treatment. The dose of anakinra suggested in our health system protocol (100mg subcutaneous four times per day for three days, followed by taper) was moderate in comparison. It is possible that the lack of benefit with anakinra may be due to lower doses and subcutaneous administration, decreasing drug availability, especially in the critically ill.

The time to treatment with immunomodulators varied from T_0_ leading us to question the role that time-to-treatment plays in modulating CCS amongst treatment arms. With anakinra, we observed a delay in drug initiation when combined with corticosteroids. Statistical analysis of drug administration variation across treatment groups was not feasible in this study due to limited availability of laboratory values and disease severity over time.

Although we were rigorous in our approach to the study design and data analysis, there are intrinsic limitations that preclude definitive conclusions in retrospective studies. The effect of systematic practice variability across the health system could not be evaluated. The use of electronic health record database limited our ability to measure other potential confounders which could have influenced provider decision making about treatments, and outcomes. Further analysis of our data is needed to evaluate the effects of immunomodulatory treatments on disease progression, including rates of thrombosis and infections.

Despite these limitations, our study is the largest to date reporting outcomes comparing the use of corticosteroids, tocilizumab, anakinra and standard of care without immunomodulatory treatment in COVID-19 CCS. Our findings suggest that, when compared to standard of care, tocilizumab and corticosteroids used together, or corticosteroids used alone, was associated with lower hospital mortality. Further investigation into the effects of differential immunomodulatory dosing, and head-to-head comparison of tocilizumab plus corticosteroids versus corticosteroids alone is warranted.

## Data Availability

Data will be made available as needed

## DECLARATIONS

### Northwell COVID-19 Research Consortium Authors

Stuart L. Cohen, MD; Jennifer Cookingham, MHA; David A. Hirschwerk, MD; Naomi I. Maria, PhD; Sanjaya K Satapathy, MD; Cristina Sison, PhD; Matthew Taylor, MD; Michael Qiu, MD, PhD

### Affiliations of The Northwell COVID-19 Research Consortium Authors

Biostatistics Unit, The Feinstein Institute for Medical Research, Northwell Health, Manhasset, NY (Sison); Department of Information Services, Northwell Health, New Hyde Park, New York (Qiu); Division of Critical Care Medicine, Cohen Children’s Medical Center of New York, Northwell Health, New Hyde Park, NY and the Institute of Cancer Research, Feinstein Institutes for Medical Research (Taylor); Divisions of Hepatology, Department of Medicine, Northwell Health, Manhasset, NY (Satapathy); Donald and Barbara Zucker School of Medicine at Hofstra/Northwell, Northwell Health, Hempstead, New York (Cohen, Hirschwerk, Satapathy); Institute of Health Innovations and Outcomes Research, Feinstein Institutes for Medical Research, Northwell Health, Manhasset, New York (Cohen, Cookingham); Institute of Molecular Medicine, The Feinstein Institutes for Medical Research, Manhasset, NY (Maria)

### Disclaimer

The initial characteristics of 5700 patients from Northwell are presented elsewhere.^13^ This case series presented in-depth results on the clinical status of patients treated with corticosteroids, tocilizumab and/or anakinra that was not presented in that article.

### Author’s contributions

*Concept, design, drafting, and editing of the manuscript:* S.N., D.G.S., A.S.C., A.G.W., G.M., B.K., P.M., O.B., A.L., M.L.L. and N.H. *Editing of manuscript:* S.L.C, J.C., D.A.H., N.I.M., S.K.S., C. S., M.T. and M.Q.

#### Acknowledgements

The authors thank the Northwell COVID-19 Research Consortium for facilitating the study.

